# Predicting the response to Neoadjuvant Chemotherapy. Can the addition of tomosynthesis improve the accuracy of CESM? A comparison with breast MRI

**DOI:** 10.1101/2022.08.26.22279254

**Authors:** Sarah L Savaridas, Sarah Vinnicombe, Violet Warwick, Andrew J Evans

## Abstract

**Background:** Neoadjuvant chemotherapy (NACT) is used to downstage breast cancer prior to surgery. Image monitoring is essential to guide treatment and to assess *in vivo* chemosensitivity. Breast MRI is considered the gold-standard imaging technique; however, it is contraindicated or poorly tolerated in some patients and may be hard to access. Evidence suggests contrast enhanced spectral mammography (CESM) may approach the accuracy of MRI. This novel pilot study investigates whether the addition of digital breast tomosynthesis (DBT) to CESM increases the accuracy of response prediction.

**Results:** Sixteen cancers in fourteen patients were imaged with CESM+DBT and MRI following completion of NACT. Ten cancers demonstrated pathological complete response (pCR) defined as absence of residual invasive disease. Greatest accuracy for predicting pCR was with CESM contrast-enhancement only (accuracy 81.3%, sensitivity 100%, specificity 57.1%), followed by MRI (accuracy 62.5%, sensitivity 44.4%, specificity 85.7%). Concordance with invasive tumour size was greater for CESM than MRI, concordance-coefficients 0.70 vs 0.66 respectively. MRI demonstrated greatest concordance with whole tumour size followed by CESM contrast-enhancement plus microcalcification, concordance-coefficients 0.86 vs 0.69. The addition of DBT did not improve accuracy for prediction of pCR or residual disease size. Whereas CESM+DBT tended to underestimate size of residual disease, MRI tended to overestimate but no significant differences were seen (p>0.05).

**Conclusions:** CESM contrast-enhancement plus microcalcification is similar to MRI for predicting residual disease post-NACT. Size of enhancement alone demonstrates best concordance with invasive disease. Inclusion of residual microcalcification improves concordance with DCIS. The addition of DBT to CESM does not improve accuracy.

**Highlights:**

- No benefit of adding DBT to CESM for NACT response prediction
- CESM appears similar to MRI for predicting response to NACT
- CESM has greatest accuracy for residual invasive tumour size.
- CESM+calcification has greater accuracy for predicting residual *in situ* disease.

## Introduction

With developments in oncological treatment, increasing numbers of women with breast cancer are receiving pre-surgical neoadjuvant chemotherapy (NACT), to downstage inoperable locally advanced disease, and to reduce the extent of surgery in both breast and axilla in women with operable disease.^1^

Imaging monitoring of treatment response is necessary during NACT to assess chemosensitivity and aid surgical decision making. Currently contrast-enhanced magnetic resonance imaging (MRI) is considered the gold-standard technique for predicting both residual tumour size and pathological complete response (pCR).^2-4^ Unfortunately, it is an expensive and time-consuming technique that may be hard to access due to service pressures. Furthermore, for some patients, it is either contraindicated or poorly tolerated.^5-7^ Whilst MRI consistently surpasses the standard imaging techniques of mammography and grey scale ultrasound for response prediction,^2^ an increasing body of evidence suggests that advanced mammographic techniques of contrast enhanced mammography (CESM) and digital breast tomosynthesis (DBT) may have comparable accuracy.^8-11^

CESM is a functional imaging technique which produces both low-energy mammograms, equivalent to full-field digital mammography, and a reconstructed image which demonstrates the vascularity of breast lesions through dual energy subtraction. DBT is a pseudo-3D mammographic technique, which eliminates overlapping breast tissue and improves visibility of malignant structural features, particularly spiculation, with increased cancer detection rates, especially in dense breasts, when compared with 2D mammograms.^12^ Recent technological developments allow a DBT acquisition during the same breast compression as a CESM study.

In this novel pilot study, we hypothesised that the addition of CESM to DBT may improve accuracy by combining the functional data of CESM with the morphological information derived from DBT. Unlike in previous research, the step-wise additional benefit of the low-energy mammogram, DBT and subtracted CESM image are considered in comparison with MRI for prediction of response to NACT.

## Methods

This was an ethically-approved prospective, paired imaging comparison study: CONtrast enhanced Digital breast tomosynthesis for monitoring Of Response to neoadjuvant chemotherapy (CONDOR), researchregistry5895. Women aged over 18 years with invasive cancers undergoing NACT were eligible for inclusion. Exclusion criteria were contraindication to iodinated contrast, contraindication to MRI, history of previous breast cancer surgery or implants, and current pregnancy or lactation. Study participants were imaged using CESM+DBT alongside standard-of-care MRI prior to NACT and at the end of NACT. Our standard protocol consisted of six cycles of FEC-T [fluorouracil (5FU), epirubicin, cyclophosphamide and docetaxel]. Chemotherapy regimens were modified in cases of co-morbidity/frailty and drug reactions.

### CESM+DBT protocol

CESM+DBT images were acquired using the Selenia Dimensions system (Hologic™, Massachusetts, USA). Imaging was commenced 3 minutes after intravenous administration of 1.5mg/kg iodinated contrast agent (Omnipaque 300, GE Healthcare™, Buckinghamshire, UK), at a rate of 2-3ml/second. After 3 minutes, imaging was commenced, consisting of bilateral craniocaudal and oblique views prior to NACT and two view ipsilateral examinations at the end of treatment.

### MRI protocol

Breast MRI was performed on a Siemens 3T Prisma Fit scanner (Siemens Healthineers, Erlangen, Germany), using a dynamic contrast-enhanced protocol. The sequences included T1 2D axial high resolution, T2 axial turbo spin echo, diffusion sequences, T1 3D dynamic sequences (2 pre-contrast and 7 post-contrast) and a delayed T1 axial high-resolution sequence, with a total scan time of approximately 40 minutes.

### Histopathology

Histology data were recorded from the diagnostic core biopsy and surgical excision specimen. Grade, tumour type and receptor status were assessed on the core biopsy specimen while residual whole tumour size (WTS) and invasive tumour size (ITS) were assessed on the resection specimen. Pathological complete response was defined as the absence of residual invasive disease within the breast (ypT0/is).^13^

### Measurement of response

All imaging assessment by readers was blinded to pathological outcomes. Patients with matched CESM+DBT and MRI end-of-treatment imaging were included, and maximum suspicious disease dimensions were recorded in each affected breast. All components of CESM+DBT were read in sequence - low energy (LE) mammogram followed by DBT then CESM – therefore the LE mammogram was read with no prior imaging while the CESM was read knowing what the mammogram and DBT had shown. The size and location of lesion(s), and total suspicious disease extent was recorded for each. CESM+DBT images were reported by a breast radiologist blinded to the MRI findings.

Similarly, MRI scans were read by an experienced radiologist blinded to CESM+DBT findings but aware of the LE mammogram findings. Lesion position, size and total disease extent was documented. Resolution of mass or malignant microcalcification was considered a complete imaging response on LE mammogram and DBT. No enhancement above background was considered a complete response on CESM and MRI. To assess the additive benefit of the CESM+DBT study, two further components were considered; CESM+calc – the maximum dimension of enhancement and/or mammographic microcalcification, and CE-DBT – the maximum area of enhancement, mammographic microcalcification and/or DBT abnormality.

Pathological results, ITS and WTS were considered the ‘ground truth’. Analysis was conducted at lesion level. In cases of pathological multifocality, the size of individual lesions was considered separately. Concordance of residual WTS and ITS with size of residual disease as predicted by each imaging modality was assessed. Both the signed difference - where a negative value indicates an imaging underestimate of pathological size and a positive value indicates an overestimate, and the absolute difference were recorded. For prediction of pCR, analysis was conducted at ‘breast level’. For patients with bilateral cancers the response in each breast was considered separately.

### Statistical analysis

Sensitivity was defined as the proportion of lesions demonstrating pCR at surgical excision with a corresponding imaging complete response; and specificity the proportion of lesions with residual invasive disease (non-pCR) with an incomplete response on imaging.^2^ Concordance of residual WTS and ITS with size of residual disease as predicted on each imaging modality was calculated using Lins concordance coefficient.^14^ Difference between MRI size and pathology size vs components of CESM+DBT and pathology was calculated using the T-test for dependent means, p<0.05 was taken as the limit of statistical significance. Statistical analyses were performed using SPSS (SPSS for Windows. 2017, v25. Armonk. NY: IBM Corp) and MedCalc (MedCalc for Windows, v20.011). Ostend, Belgium: MedCalc Software). Software was chosen according to availability of required functionality.

## Results

Eighteen of 31 (58%) eligible patients were recruited. Three patients could not be recruited due to logistical issues regarding availability of pre-treatment CESM+DBT after the decision to treat with NACT and the first chemotherapy cycle. Average participant age of participants was 52.7 years (range 32-72 years). Fourteen patients received FEC-T chemotherapy, two patients FEC-only chemotherapy (6 cycles) and two, taxane-only chemotherapy (4 cycles) due to comorbidities.

### Histopathology

Multifocal disease was present in five cases. Three women had unilateral multifocal disease (two tumours), two had bilateral disease. One of the women with bilateral disease had three distinct tumours. In total, there were 24 invasive carcinomas. One was a mammographically occult invasive lobular cancer (ILC), in a patient with bilateral invasive ductal carcinoma (IDC), the remainder were IDC. With respect to invasive tumour grade, fourteen (58%) were grade 3, nine (38%) grade 2 and one (4%) grade 1. Regarding receptor status, eleven (46%) were ER/PR+ve HER-ve, ten (42%) were HER2+ve and three (13%) were triple negative.

### Imaging Protocol

There were no significant adverse events. One patient withdrew at mid-treatment because of difficult intravenous access. One patient developed bone metastases and treatment became palliative. The two patients who had 4 cycles of taxane-only chemotherapy did not have end-of-treatment MRI as per local hospital guidelines. Therefore, 14 patients (16 breasts) had both CESM+DBT and MRI at end-of-treatment. There was no significant difference in the mean interval between imaging and surgery; 25.71 days, (range:13-42) and 25.79 days (range:19-38) for CESM+DBT and MRI respectively, *p*=0.711.

### Prediction of pCR on post-chemotherapy images

Of the 16 breasts with cancer, ten demonstrated a pathological response (pCR), prevalence 62.5%. The diagnostic accuracy of each imaging modality for predicting pCR is illustrated in table 1.

**Table 1:**
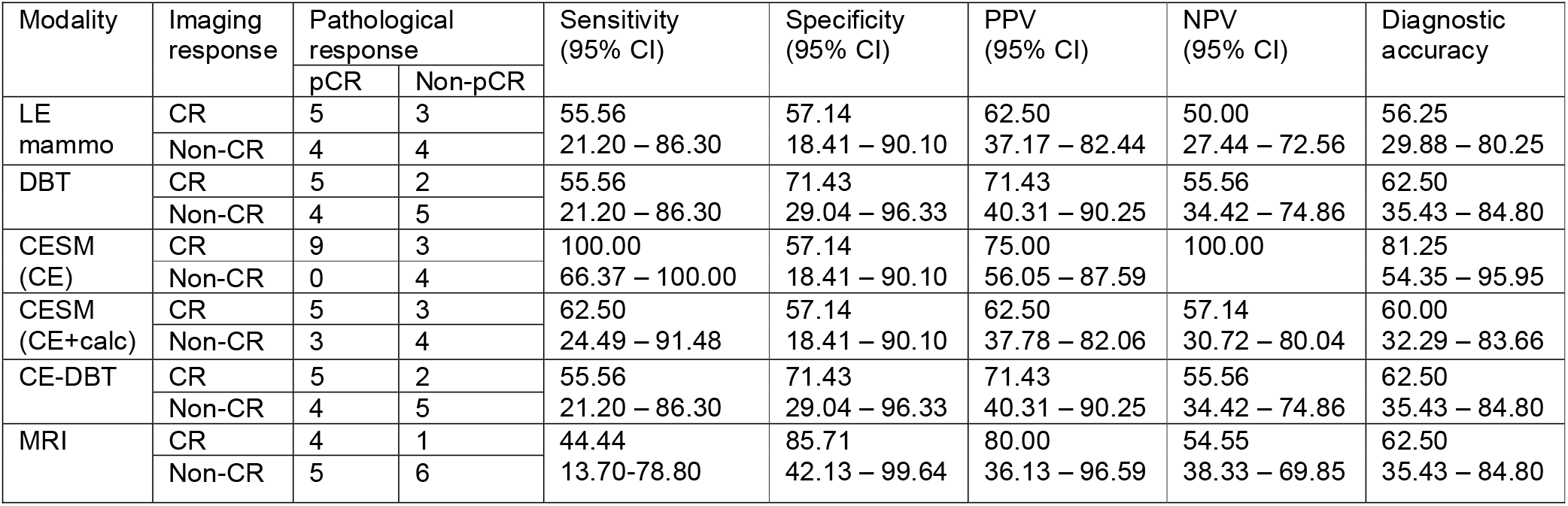
Diagnostic accuracy for predicting pCR according to imaging technique

The greatest accuracy for predicting pCR, 81.25%, was seen with CESM (corresponding sensitivity for MRI only 55.6%). All patients with pCR were identified. However, three patients were incorrectly classified as complete responders due to lack of residual enhancement. In two of these breasts the foci of residual invasive disease measured 6mm or less. In the third case whilst there was ‘marked and almost pathological complete response to neoadjuvant chemotherapy… more than 90% loss of tumour cellularity’, foci of invasive disease persisted over an area of 72mm. The same residual disease was also occult on DBT in two cases, and for all three with mammography. Sensitivity for pCR was lower for mammography and DBT. The combined measure of CESM+calc resulted in lowered sensitivity with no improvement in specificity. CE-DBT demonstrated an incremental increase in specificity but larger drop in sensitivity compared to CESM.

MRI had the highest specificity with only one false positive – a 6mm site of invasive disease. However, MRI only identified 5 patients with pCR, resulting in a lower sensitivity (55.6%) and lower overall diagnostic accuracy.

### Prediction of residual tumour size on post-chemotherapy imaging

The individual components of CE-DBT demonstrate similar reliability for predicting WTS, with concordance coefficients for mammography, DBT and CESM of 0.68, 0.65 and 0.53 respectively. The combined assessment CESM+calc increased concordance to 0.70. No benefit was seen when combining with DBT, with an overall CE-DBT concordance of 0.67. MRI conferred the strongest concordance (0.87). By comparison CESM and MRI confer similar concordance for predicting ITS, concordance coefficients 0.70 and 0.66 respectively.

Both signed and absolute difference between imaging size and pathology are displayed in table 2.

**Table 2:**
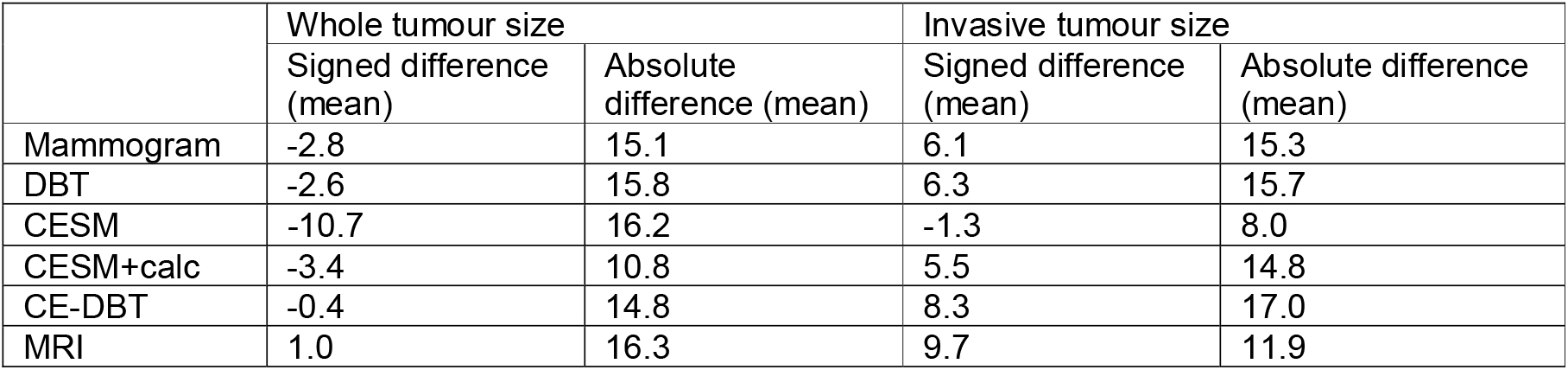
Signed and absolute difference between imaging size and pathological size

|  | Whole tumour size |  | Invasive tumour size |  |
| --- | --- | --- | --- | --- |
|  | Signed difference (mean) | Absolute difference (mean) | Signed difference (mean) | Absolute difference (mean) |
| Mammogram | -2.8 | 15.1 | 6.1 | 15.3 |
| DBT | -2.6 | 15.8 | 6.3 | 15.7 |
| CESM | -10.7 | 16.2 | -1.3 | 8.0 |
| CESM+calc | -3.4 | 10.8 | 5.5 | 14.8 |
| CE-DBT | -0.4 | 14.8 | 8.3 | 17.0 |
| MRI | 1.0 | 16.3 | 9.7 | 11.9 |

The signed differences indicate that all components of CESM+DBT tend to underestimate whole tumour size with CESM demonstrating a mean underestimation of 10.7mm which is reduced to 3.4mm and 0.4mm when combined with the presence of residual microcalcification and DBT findings. By comparison, MRI overestimates whole tumour size by an average of 1mm. However, when the absolute difference is considered, CESM+calc demonstrates the closest estimation of 10.8mm, with a mean difference of 16.3mm for MRI.

For invasive tumour size the signed mean indicates an underestimation of only 1.3mm for CESM, with an absolute difference of 8mm. By comparison MRI tends to overestimate invasive disease extent by an average of 9.7mm (signed difference) and 11.9mm (absolute difference). No significant difference was seen between MRI and all other modalities for signed or absolute differences, *p* > 0.05.

## Discussion

We have demonstrated that the use of CE-DBT for monitoring response to NACT within clinical care is feasible. Of those patients meeting inclusion criteria, 58% were recruited. This may have increased to 68% had it been possible to offer timely pre-treatment CE-DBT to an additional three patients. With regard to a future multi-centre trial, it is likely that enrolment would be higher in centres using CESM+DBT at time of diagnosis. There were no adverse outcomes reported during the trial, one patient withdrawing at mid-treatment because of poor intravenous access. Of note, the challenges around recruitment would not apply if this modality were introduced as routine clinical practice.

Consistent with previous studies we have shown that CESM has greater accuracy at predicting pCR than mammography.^10,15-18^ With respect to DBT, our results are similar to two studies which compared DBT to mammography and ultrasound, reporting a sensitivity of 44.7-50% and specificity of 91-97.6%.^11,19^ However this is the first study to consider the combined use of CESM and DBT in the context of NACT. We have demonstrated lower accuracy for DBT than CESM with no additive value in combining DBT with CESM. Thus, our findings do not support the combined use of CESM+DBT as a modality for detection of pCR, especially when the increased radiation dose is taken into consideration.

With regard to MRI, whilst we demonstrate a similar specificity and accuracy to previous studies comparing CESM and MRI, our sensitivity is lower.^8,9,15^ This may be related to variation of pCR definition. Indeed, meta-analysis of MRI studies demonstrated that those that permitted residual DCIS in the definition of pCR – such as our study – tended to demonstrate lower accuracy, AUC 0.83 vs 0.88.^18^

In addition to predicting pCR pre-operatively, it is important to quantify the size of residual disease to guide surgery – whether breast conserving surgery is feasible, to improve surgical margins and reduce surgical re-excision rates. Whilst presence of residual *in situ* disease in the absence of invasive disease does not affect survival or local recurrence rate,^13^ it is important for surgical decision making. Therefore, we considered both residual whole tumour size (WTS) for surgical decision making, as well as invasive tumour size (ITS) for prognostication.

In our study, CESM enhancement demonstrated the greatest accuracy for predicting residual ITS, the greatest concordance occurring with CESM enhancement alone, followed by MRI. Regarding WTS, MRI demonstrated the greatest concordance with promising results seen for CESM, especially when microcalcification was considered in addition to residual enhancement. No significant difference was seen between the accuracy of MRI and CESM+DBT.

Our results are consistent with published data on CESM for the prediction of residual disease, with concordance coefficients ranging from moderate to good, 0.7-0.81.^8,9,15^. Our findings are consistent with those of Iotti *et al* who report that the addition of a measurement of microcalcification to the diameter of residual enhancement increases sensitivity for detection and accurate measurement of residual disease, though it reduces specificity.^20^ Furthermore, it is accepted that the presence of residual mammographic microcalcifications is not consistently related to residual disease, and that even with loss of MRI enhancement it is not possible to predict absence of residual disease with sufficient accuracy to avoid complete excision of tumour bed calcifications.^21-23^ We suggest that this finding is also true for persistent microcalcifications in the absence of CESM enhancement.

Our results for DBT and residual tumour size assessment are consistent with the limited published literature. Park *et al* reported an intraclass correlation coefficient of 0.63, with mean difference between DBT and pathology of 16.6mm^11^.

This is a novel exploratory study and thus, was not powered to detect significant differences in the performance of CE-DBT and MRI. However, importantly we demonstrated no benefit from incorporating DBT to produce a full CE-DBT score for predicting either WTS or ITS.

The main limitation of this study is the small numbers of patients. Although this is partially mitigated by the fact that it is a prospective study allowing direct comparison of two imaging techniques, it is acknowledged that this limits the weight that can be given to the statistical analysis. No assessment of inter-reader reproducibility was possible as the CESM+DBT and MRI studies were each interpreted by single but independent readers. However, we have demonstrated comparable accuracy for CESM+DBT studies interpreted by a relatively inexperienced reader, compared to MRI studies reported by an expert with extensive MRI experience. The small numbers preclude evaluation of performance by imaging phenotype and tumour subtype, and further research into this is needed.

## Conclusions

The findings of this pilot study do not support the addition of DBT to CESM for detecting pCR or size of residual disease following NACT. We suggest CESM is similar to MRI for predicting pCR and residual invasive tumour size. We recommend that the residual contrast enhancement on recombined CESM images is reported in parallel with residual microcalcifications on the low energy mammograms to improve accuracy of predicting residual *in situ* disease.

## Data Availability

All data produced in the present study are available upon reasonable request to the authors

## List of abbreviations

CESM: Contrast-enhanced spectral mammography
DCIS: Ductal carcinoma in situ
DBT: Digital breast tomosynthesis
FEC-T: Fluorouracil (5FU), epirubicin, cyclophosphamide and docetaxel
FFDM: Full field digital mammography
IDC: Invasive ductal carcinoma
ILC: Invasive lobular carcinoma
ITS: Invasive tumour size
MRI: Magnetic resonance imaging
NACT: Neoadjuvant chemotherapy
pCR: Pathological complete response
WTS: Whole tumour size

## Notes

### Competing Interest Statement

The authors have declared no competing interest.

### Clinical Trial

researchregistry5895

### Funding Statement

TENOVUS Scotland
British Society of Breast Radiologists

### Author Declarations

Research ethics committee of the West of Scotland gave ethical approval for this work

